# Reassessment of persistent symptoms, self-reported COVID-19 infection and SARS-CoV-2 serology in the SAPRIS-SERO cohort: identifying possible sub-syndromes of Long Covid

**DOI:** 10.1101/2022.02.25.22271499

**Authors:** Nicola Spiers

## Abstract

**Background:** Long Covid remains a relatively new phenomenon with emerging understanding. Estimated UK prevalence of Long Covid with three or more symptoms lasting for 12 weeks or more was 2.2% at the end of 2021. The population-based French SAPRIS-SERO cohort has novel information about the pattern of symptoms of Long Covid that has been obscured by controversy around the original paper.

**Methods:** Secondary analysis was used to describe and re-interpret the frequencies of persistent symptoms by IgG seropositivity and self-reported Long Covid in the published report of the SAPRIS-SERO survey. Participants in the cross-sectional analysis were 26 823 individuals from the French population-based CONSTANCES cohort, included between 2012 and 2019, who took part in the nested SAPRIS and SAPRIS-SERO surveys. Between May and November 2020, the Euroimmun enzyme-linked immunosorbent assay was used to detect anti-SARS-CoV-2 antibodies. Surveyed online between December 2020 and January 2021, participants self-reported previous COVID-19 infection and physical symptoms during the previous four weeks that were new since March 2020, and had persisted for at least eight weeks.

**Results:** There was similarity of prevalence over the majority of symptoms in those self-reporting COVID-19 infection, regardless of blood test result. Persistent symptoms significantly associated with self-reported COVID-19 infection and common in both groups include respiratory tract symptoms and a group of symptoms that might be related to chronic fatigue, malaise or postural issues. Seropositivity for IgG antibodies did not predict symptoms independently of self-reported Long Covid, except for anosmia.

**Conclusions:** There may be three common sub-syndromes of Long Covid, one with persistent anosmia, another with other respiratory tract symptoms and a third, currently under researched, with symptoms relatable to chronic fatigue. Antibody tests are insufficient for case detection while Long Covid remains poorly understood.

**Key Messages:**

- IgG seropositivity is insufficient to identify potential cases of Long Covid
- Persistent anosmia is very strongly associated with IgG seropositivity and may define a subsyndrome of Long Covid
- Other potential subsyndromes are those with persistent respiratory symptoms and those with persistent symptoms relatable to fatigue, malaise or postural issues
- The Long Covid research effort should be rebalanced towards understanding the fatigue/tiredness subsyndrome.

## Introduction

Long Covid remains a relatively new phenomenon with emerging understanding^1^. Estimated UK prevalence of Long Covid with three or more symptoms lasting for 12 weeks or more was 2.2% at the end of 2021^2^, while prevalence of symptoms lasting for 12 weeks or more and daily activities limited a lot was 0.36%^3^. Population-based studies of Long Covid that describe persistent symptoms after COVID-19 infection are rare.

Associations of persistent symptoms with SARS-CoV-2 antibodies and self-reported COVID-19 infection in the SAPRIS-SERO survey were first reported in November 2021^4^. The authors’ conclusion, that persistent physical symptoms after COVID-19 infection may be associated more with belief in having been infected than with laboratory confirmed infection, has proved controversial. The controversy has overshadowed novel evidence from SAPRIS-SERO on the pattern of persistent symptoms of Long Covid. This paper presents a secondary analysis of symptom frequencies from the original paper to support reassessment of findings.

## Methods

Participant enrolment and measurements are described in detail in the original paper^4^. Participants responding to an annual questionnaire from the CONSTANCES population^5^ were invited to complete the Euroimmun assay for SARS-CoV-2 IgG antibodies between May and November 2020 and an internet questionnaire between December 2020 and January 2021. Participants were asked “Since March, do you think you have been infected by the coronavirus (whether or not confirmed by a physician or test)?”, being aware of their blood test result at the time of the interview. Participants reported a list of 18 new persistent symptoms since March 2020 that were present in the last 4 weeks and had persisted for more than 8 weeks.

Secondary analysis of symptom frequencies presented in Table 2 of the original paper was carried out by calculating odds ratios and Wald statistics^6^. Individual-level data was not accessed. The following analysis decisions were made a priori: Firstly, the analysis should be descriptive and form a basis for hypothesis generation. Secondly the ‘healthy’ subgroup who were seronegative and did not self-report COVID-19 infection should form the base for comparisons. Thirdly, that the seropositives who self-reported COVID-19 would provide the strongest evidence for persistent symptoms of Long Covid. For clarity, symptoms were ordered by the strength of association for this group. Finally, that language describing the results should be precise and accessible, where possible, across clinical disciplines and lay perspectives.

## Results

There were 26,823 participants with complete data. Among the majority who were seronegative and did not self-report COVID-19 infection (base; n=25271), common new persistent symptoms since March 2020 included sleep problems, musculoskeletal issues, fatigue, problems with attention or concentration, digestion and skin problems (Table). The seropositives who did not self-report COVID-19 infection, despite being aware of their positive antibody test (n=638), had similar symptom prevalence to the base group. Exceptions were anosmia, based on small numbers, and skin problems and joint pain, where prevalence was lower.

**Table 1.** New persistent symptoms by self-reported COVID-19 and antibody test status

| Self-reported COVID-19<br>IgG Antibody test | Not infected |  | Infected |  |
| --- | --- | --- | --- | --- |
|  | Negative | Positive | Negative | Positive |
|  | n=25271 | n=638 | n=461 | n=453 |
|  | % base | % | % | % |
| anosmia | 0.3 | 1.1*** | 4.3*** | 9.9*** |
| chest pain | 0.5 | 0.3 | 3.0*** | 4.4*** |
| breathing difficulties | 0.8 | 1.4 | 6.3*** | 5.7*** |
| fatigue | 2.5 | 3.4 | 12.4*** | 13.7*** |
| palpitations | 0.7 | 0.9 | 3.7*** | 3.3*** |
| cough | 0.6 | 0.3 | 2.2*** | 2.4*** |
| cognitive<sup>a</sup> | 2.2 | 2.7 | 7.4*** | 8.4*** |
| other | 1.8 | 1.3 | 3.7** | 5.7*** |
| dizziness | 0.6 | 0.8 | 1.5* | 1.8** |
| headache | 1.3 | 1.3 | 2.8** | 3.5*** |
| back pain | 6 | 5.2 | 6.9 | 8.8* |
| muscular pain/soreness | 3.2 | 2.8 | 4.8 | 4.2 |
| hearing | 1.8 | 0.9 | 1.5 | 2.2 |
| digestive tract problems | 3.3 | 3.1 | 7.2*** | 4 |
| joint pain | 7.1 | 4.1** | 6.5 | 7.9 |
| skin | 2.4 | 0.9* | 3.7 | 2.4 |
| sleep problems | 10.2 | 8.6 | 10.6 | 9.9 |
| sensory | 1.8 | 1.3 | 3.5* | 1.1 |

Odds ratio compared with base\*P<0.05 \*\*P<0.01 \*\*\*P<0.001 <sup>a</sup>poor attention/concentration

The pattern of symptoms in participants self-reporting COVID-19 infection, supported by a positive blood test (n=453) was quite different from the base. Prevalence of persistent new respiratory tract symptoms (anosmia, chest pain, breathing difficulties, cough) were all raised, as were prevalence of a second group of symptoms that can be related to chronic fatigue or postural issues (fatigue, palpitations, poor attention/concentration, dizziness, headache, back pain).

The comparison between those self-reporting COVID-19 infection but seronegative (n=461) and the base is similar, with prevalence of all four respiratory symptoms raised, and four of the five fatigue-related symptoms. Indeed, the prevalence of symptom reporting is similar in those self-reporting COVID-19, regardless of whether this is supported by a positive antibody test. Exceptions are anosmia, more strongly associated in seropositives, back pain, associated only in seropositives, and sensory symptoms and digestion, associated in seronegatives who self-reported COVID-19.

## Discussion

Common new persistent symptoms since March 2020 in those testing negative for antibodies and not self-reporting COVID-19 infection included sleep problems, musculoskeletal and digestive tract symptoms.

The 638 seropositives who did not self-report COVID-19 infection had generally similar symptom prevalence to the base group. This may be explained by false positives on the assay, given that, with sensitivity of 87% and specificity of 97.5%, approximately 680 would be expected in this dataset. This subgroup may also include those with mild or asymptomatic illness. They reported less skin problems and joint pain, giving some support to hypotheses of lower COVID-19 related anxiety or a higher threshold for symptom reporting.

In a model adjusting for self-reported COVID-19 infection^4^, only anosmia and skin problems (lower prevalence) remained predictive of symptom prevalence, reflecting the similarity of symptom reporting in those self-reporting COVID-19 infection, regardless of blood test result.

The exception of anosmia is highlighted by the authors of the original paper. Anosmia is a highly specific symptom for COVID-19, being rare in the base group, and very strongly associated with seropositivity (prevalence 4.7% vs 0.4%). The authors of the original paper describe persistent anosmia as a hallmark of COVID-19, and from this assumption build support for their a priori theory that seronegatives who self-report COVID-19 infection may have either misattributed symptoms or developed them from belief in having been infected. There is perhaps some support for this theory in the raised levels of sensory and digestive tract symptoms in seronegatives who self-reported COVID-19.

The issue rests on what can be deduced from lack of association with IgG antibodies at an unknown time from infection. Beyond this, the data say nothing about the biological and psychological mechanisms underlying the pattern of symptom reporting in those self-reporting COVID-19 infection, leaving room for alternative hypotheses that cannot be ruled out. It is worth noting that 24% of the seronegatives who self-reported infection also self-reported a positive PCR test^10^

One such is that individuals who developed persistent anosmia are a subgroup of those infected with SARS-CoV-2 who experienced an energetic immune response in the upper respiratory tract shortly after infection, correlated with subsequent high antibody levels detectable by the Euroimmun assay. A related hypothesis is that the Euroimmun assay failed to detect other common presentations of COVID-19 infection resulting in persistent symptoms. A form of infection that is initially mild but results in long-term fatigue and related symptoms may be associated with weak immune response and hence negative findings on the assay.

Deciding between these competing accounts will depend on progress in understanding the dynamics of SARS-CoV-2 immune response, and better understanding of COVID-19 persistent symptoms, perhaps with particular emphasis on fatigue.

As well as false positives and false negatives on the assay, already discussed, limitations of the SAPRIS-SERO cohort include ageing of the sample and selective attrition by education, income and self-rated health. The list of symptoms reported is also limited.

With REACT, SAPRIS-SERO is one of only two large-scale population-based studies, including those non-infected, of persistent symptoms after COVID-19. These provide an essential complement to non-probability surveys, positive test cohorts and numerous smaller-scale studies of Long Covid, often in hospitalised patients^7^, especially as Long Covid is often reported after mild initial disease^8^.

Although clustering of symptoms was not analysed, the SAPRIS-SERO findings are suggestive of three clusters: one with anosmia, a second with other respiratory tract symptoms: chest pain, breathing difficulties and cough, and a third with symptoms that might be related to chronic fatigue, malaise or postural issues. These findings are consistent with those reported in a preprint from REACT^2^. Symptoms persisting for 12 weeks identified by REACT fell into two clusters, a “tiredness cluster” experiencing high prevalence of tiredness, co-occurring with muscle aches, difficulty sleeping and shortness of breath, and a “respiratory cluster” experiencing high prevalence of respiratory symptoms including shortness of breath, tight chest and chest pain. Similar associations of persistent symptoms with test positivity were also reported in a convenience sample reweighted to represent the US population^9^.

These findings are timely with the onset of the Omicron variant. While it seems likely that incidence of the respiratory subsyndrome will be much lower from Omicron, incidence of the fatigue/tiredness subsyndrome, found to be three times more common than the respiratory cluster by REACT, remains unknown. It is therefore too early to conclude how far incidence of Long Covid will be reduced in the era of Omicron.

Much of the research effort on Long Covid to date has been in hospitalised cohorts, where patients who go on to experience persistent respiratory symptoms are well-represented. This is unsurprising, given that infrastructure for such research is well-developed, and the respiratory and cardiovascular systems are relatively well-understood. On the other hand, a tiredness or fatigue based subsyndrome of Long Covid presents challenges of case identification, choice of comparisons, developing measurements, and working with sufferers and across disciplines on an under researched phenomenon where there is risk of being wrong. The findings here and from REACT emphasise the urgency of taking up these challenges and rebalancing the Long Covid research effort towards understanding the tiredness cluster, and how these persistent and disabling symptoms may result from initially mild disease.

## Conclusion

Antibody tests are insufficient for case detection while Long Covid remains poorly understood. There may be three common sub-syndromes of Long Covid, one with persistent anosmia, another with other persistent respiratory tract symptoms, and a third with symptoms relatable to chronic fatigue that is currently under-researched.

## Data Availability

Source data are openly available in Table 2 of reference 4

https://jamanetwork.com/journals/jamainternalmedicine/fullarticle/2785832

## Data Availability

Source data are openly available in Table 2 of reference 4

https://jamanetwork.com/journals/jamainternalmedicine/fullarticle/2785832

## Data Availability

Source data are openly available in Table 2 of reference 4

https://jamanetwork.com/journals/jamainternalmedicine/fullarticle/2785832

## Ethics Approval

Ethics approval was not required for secondary analysis of data in the public realm.

## Data Availability

Data are available in Table 2 from the original paper (reference 4).

## Acknowledgment

The author thanks Julie Gamble for assistance with manuscript preparation

